# Application of comprehensive evaluation framework to Coronavirus Disease 19 studies: A systematic review of translational aspects of artificial intelligence in health care

**DOI:** 10.1101/2023.02.23.23286374

**Authors:** Aaron Casey, Saba Ansari, Bahareh Nakisa, Blair Kelly, Pieta Brown, Paul Cooper, Imran Muhammad, Steven Livingstone, Sandeep Reddy, Ville-Petteri Makinen

## Abstract

**Background:** Despite immense progress in artificial intelligence (AI) models, there has been limited deployment in healthcare environments. The gap between potential and actual AI applications is likely due to the lack of translatability between controlled research environments (where these models are developed) and clinical environments for which the AI tools are ultimately intended.

**Objective:** We have previously developed the Translational Evaluation of Healthcare AI (TEHAI) framework to assess the translational value of AI models and to support successful transition to healthcare environments. In this study, we apply the TEHAI to COVID-19 literature in order to assess how well translational topics are covered.

**Methods:** A systematic literature search for COVID-AI studies published between December 2019-2020 resulted in 3,830 records. A subset of 102 papers that passed inclusion criteria were sampled for full review. Nine reviewers assessed the papers for translational value and collected descriptive data (each study was assessed by two reviewers). Evaluation scores and extracted data were compared by a third reviewer for resolution of discrepancies. The review process was conducted on the Covidence software platform.

**Results:** We observed a significant trend for studies to attain high scores for technical capability but low scores for the areas essential for clinical translatability. Specific questions regarding external model validation, safety, non-maleficence and service adoption received failed scores in most studies.

**Conclusions:** Using TEHAI, we identified notable gaps in how well translational topics of AI models are covered in the COVID-19 clinical sphere. These gaps in areas crucial for clinical translatability could, and should, be considered already at the model development stage to increase translatability into real COVID-19 healthcare environments.

## Introduction

The discussion about the value of Artificial Intelligence (AI) to healthcare and how AI can address healthcare delivery issues have been in place for some years now [1–3]. However, most stakeholders are eager for this discourse to move beyond theoretical or experimental confines to adoption and integration in clinical and real-world healthcare environments [1,4,5]. Recently, we have started to see some AI applications undergoing clinical trials or integration into medical devices or medical information systems [6]. Yet most AI applications in healthcare have not demonstrated improvement in clinical or healthcare outcomes [5,7]. What prevents these applications from translating their potential to clinical outcomes? Firstly, many of these AI applications are developed to demonstrate algorithmic performance or superiority rather than improvement in clinical results [8,9]. Secondly, the applications are not considered for use beyond the experimental or pilot settings [8]. This limitation means their performance does not often generalise beyond test data sets. Thirdly, even when these applications are externally validated, they are seldom integrated into existing clinical workflows, often as a result of decreased performance on the external validation [10] or low acceptance by clinicians [11]. The latter aspect means these applications remain experimental novelties rather than useful tools for clinicians. Added to these translational issues are problems with data that may lead to inaccurate results or the introduction of biases. Several studies have shown how such issues can have adverse outcomes for patients and communities [12–14]. Yet, ethical and governance safeguards are often missing in AI in healthcare applications or studies [14].

These translational issues suggest there is a need for a comprehensive framework that can support researchers, software vendors and relevant parties in systematically assessing their AI applications for their translational potential. To address this gap, we formed an international team and ran a systematic process over 18 months to develop an evaluation and guidance framework, termed ‘Translational Evaluation of Healthcare AI (TEHAI)’ [15]. This framework focuses on the aspects that can support the practical implementation and use of AI applications. TEHAI has three main domains (capability, utility, and adoption components) and 15 subcomponents (Table 1 and Multimedia Appendix 1 Eval and Scoring). As the range of clinical challenges and potential AI solutions is very wide, it is infeasible to automate the evaluation using current technology. Instead, we rely on TEHAI as an expert-driven but formalized framework where the subjectivity of an individual reviewer is mitigated by the consensus power of multiple committee members.

**Table 1.** Overview of the TEHAI framework. The framework comprises 15 separate criteria (sub-components) that are grouped into three higher-level components. Each criterion yields a score between 0 and 3 points depending on the quality of the study. To compare two or more AI models against each other, further weighting of the scores can be applied to emphasize translatability. However, in this study, the weighting was not used since we focused on the statistics of the sub-components instead.

| Component | Sub-component | Initial Score | Weight |
| --- | --- | --- | --- |
| Capability | Objective of Study | 0-3 | 10 |
|  | Dataset Source and Integrity | 0-3 | 10 |
|  | Internal Validity | 0-3 | 10 |
|  | External Validity | 0-3 | 10 |
|  | Performance Metrics | 0-3 | 10 |
|  | Use Case | 0-3 | 5 |
| Utility | Generalizability and Contextualisation | 0-3 | 10 |
|  | Safety and Quality | 0-3 | 10 |
|  | Transparency | 0-3 | 10 |
|  | Privacy | 0-3 | 10 |
|  | Non-Maleficence | 0-3 | 10 |
| Adoption | Use in a Healthcare Setting | 0-3 | 10 |
|  | Technical Integration | 0-3 | 10 |
|  | Number of Services | 0-3 | 5 |
|  | Alignment with Domain | 0-3 | 5 |

The emergence of the COVID-19 pandemic has resulted in several studies and papers outlining the utility of AI in tackling various aspects of the disease like diagnosis, treatment, and surveillance [16–19] The number of AI papers published either as pre-print or peer-reviewed has been unprecedented, even leading to the development of AI applications to keep up with and summarise the findings for scientists [20]. Some recent reviews have outlined how most of these studies or the AI applications presented in these studies have shown minimal value for clinical care [7,21]. This finding aligns with the discussion about the translational problem of AI in healthcare.

The aim of this study is to assess the awareness and consideration for important translational factors in scientific literature related to COVID-19 machine learning applications. We chose the narrow scope to ensure that our method of evaluation (i.e. TEHAI) would not be confounded by the differences that are inherent to any particular area of health care. For the reason, we included only studies where AI was clearly aimed at solving a practical problem rather than discovering new biology or novel treatments. This cost-effective approach enables us to uncover the translational gaps of the AI applications and validate the usefulness of a variety of AI models without the added complexity from high diversity of diseases or health care challenges.

## Methods

### Data extraction

Eligible studies included those where a statistical algorithm was applied to or trained with a COVID-19 dataset and where the intended use of the algorithm was to address a COVID-19 healthcare problem. Excluded studies included those where participants were younger than 18 years old and where the full text of the study was not in English. To find papers eligible for this study we searched the NIH iSearch COVID-19 portfolio, MEDLINE via Ovid, and Embase via Embase.com. These sources were searched on 7 December 2020 using search strategies consisting of keywords expected to appear in the title or abstract of eligible studies, and index terms specific to each database except in the case of the NIH iSearch COVID-19 portfolio. The search strategy was developed by a health librarian (B.K.) in consultation with the rest of the research team.

For the COVID-19 element of the search, we adapted the Wolters Kluwer expert search for COVID-19 on MEDLINE. Specifically, we removed the search lines for excluding non-COVID-19 coronaviruses (e.g., Middle East respiratory syndrome) and for pharmaceutical treatment options (e.g., Remdesivir); at the time our search strategy was created these were lines five and nine, respectively, in the Wolters Kluwer OVID COVID-19 expert search. For the AI element of the search, we searched MEDLINE for relevant papers, recording significant keywords from their titles and abstract. We also searched the Medical Subject Headings (MeSH) thesaurus for related MeSH terms. These steps led to the creation of a draft search strategy which was then tested and finalised. The search was limited to records with a publication date of 1 December 2019 onwards. This limit was to reduce the number of irrelevant results, given that the first known case of COVID-19 occurred in December 2019 (Multimedia appendix 2 Search Strategies).

A foundational Ovid MEDLINE search strategy was then translated for Embase.com to make use of appropriate syntax and index terms (Multimedia appendix 2). Similar translation was done for the NIH iSearch COVID-19 portfolio except for index terms as this resource did not use indexing at the time of search development (Multimedia appendix 2). Finally, search strategy validation and refinement took place by testing a set of known relevant papers against the search strategy as developed, with all papers subsequently recalled by the search in MEDLINE and Embase. A full reproduction of the search strategies for each database can be found in Multimedia appendix 2. Searching these databases using the search strategy resulted in 5,276 records. After removal of duplicates, we screened 3,830 records for relevance. This resulted in 968 studies identified as relevant and eligible for evaluation. From these, a sample of 123 was randomly selected for evaluation and data extraction of which 102 were included in the final set. Our target number for full evaluation was 100, however, additional papers were randomly picked to account for the rejection of 19 papers that passed the initial screen, but were deemed ineligible after closer inspection. Early on in the evaluation it became apparent that a significant portion of the studies focused on image analysis, we then enriched the pool for studies that were not image focused taking the ratio of image:non-image focused studies to 1:1. Full text was retrieved for all 123 in the randomised sample, however only 102 studies met our inclusion criteria at the evaluation and extraction stage (Multimedia appendix 4). Of the studies that did not meet our inclusion criteria, the majority were non-imaging studies and the final ratio of imaging:non-imaging focused studies was 2:1.

Evaluation and data extraction was conducted using Covidence systematic review software [22]. We used this software to facilitate the creation of a quality assessment template based on the TEHAI framework [15] in combination with other questions (henceforth referred to as data extraction questions) aimed at further understanding the components that may influence a studies capacity to translate into clinical practice (Multimedia Appendix 3). As a measure to minimize the impact of subjectivity introduced by human evaluation, each paper was initially scored by two reviewers who independently evaluated the paper against the elements of the TEHAI framework and extracted relevant data. A third reviewer then checked the scores and if discrepancies were present, chose one of the two independent reviewers’ scores as the final result. This process was built-in to the Covidence platform. To further minimize the impact of subjectivity introduced by human evaluation, reviewer roles were also randomly assigned across the evaluation team.

For scoring of the included studies, we derived upon previously provided guidance for scoring evidence within a TEHAI framework [15]. The TEHAI framework is composed of three overarching components: capability, utility and adoption. Each component numerous sub-component questions of which there are 15 in total. Scoring of each TEHAI subcomponent is based on a range of zero to three depending on the criteria met by the study. In this study, we also investigate the sums of these scores at the component level to provide a better overview of data. In addition, TEHAI facilitates direct comparisons between specific studies by a weighting mechanism that further emphasizes the importance of translatability (see the last column in Table 1). However, for the purpose of this study, where we focus on the aggregate statistical patterns, the weighting was not used.

We also asked reviewers to report on a select number of data extraction questions that would enable us to further tease apart which components of a study may influence the score obtained. These questions covered 1) the broad type of the AI algorithm, 2) methodological or clinical focus, 3) open source or proprietary software, 4) dataset size, 5) country of origin, and 6) imaging or non-imaging data.

### Data Analysis

Associations between groupings of papers and the distributions of sub-component scores were assessed by the Fisher’s exact test. Correlations between sub-components were calculated using Kendall’s formula. Component scores were calculated by adding the relevant sub-component scores together; group differences in mean component scores were assessed by the t-test. As there are 15 sub-components, we set a multiple testing threshold of *P* < 0.0033 to indicate 5% type 1 error probability under the Bonferroni correction for 15 independent tests. Unless otherwise indicated mean scores were calculated ± standard error.

## Results

### TEHAI sub-component scores

Nine reviewers reviewed a total of 102 manuscripts (mean = 22.67 per reviewer, SD = 7.71, min = 11, max = 36), with the same two reviewers scoring the same manuscript an average of 2.83 times (SD = 2.58, min = 0 max = 13). The Cohen’s kappa statistic for inter-reviewer reliability was κ = 0.45 with an asymptomatic standard error of 0.017 over the two independent reviewers. Overall, the capability component scored the highest mean score, followed by adoption and utility (Figure 1A). At the subcomponent level, the poorest performing questions were non-maleficence (93/102 scoring zero points), followed closely by safety and quality, external validity and number of services (Figure 1B).

**Figure 1.**
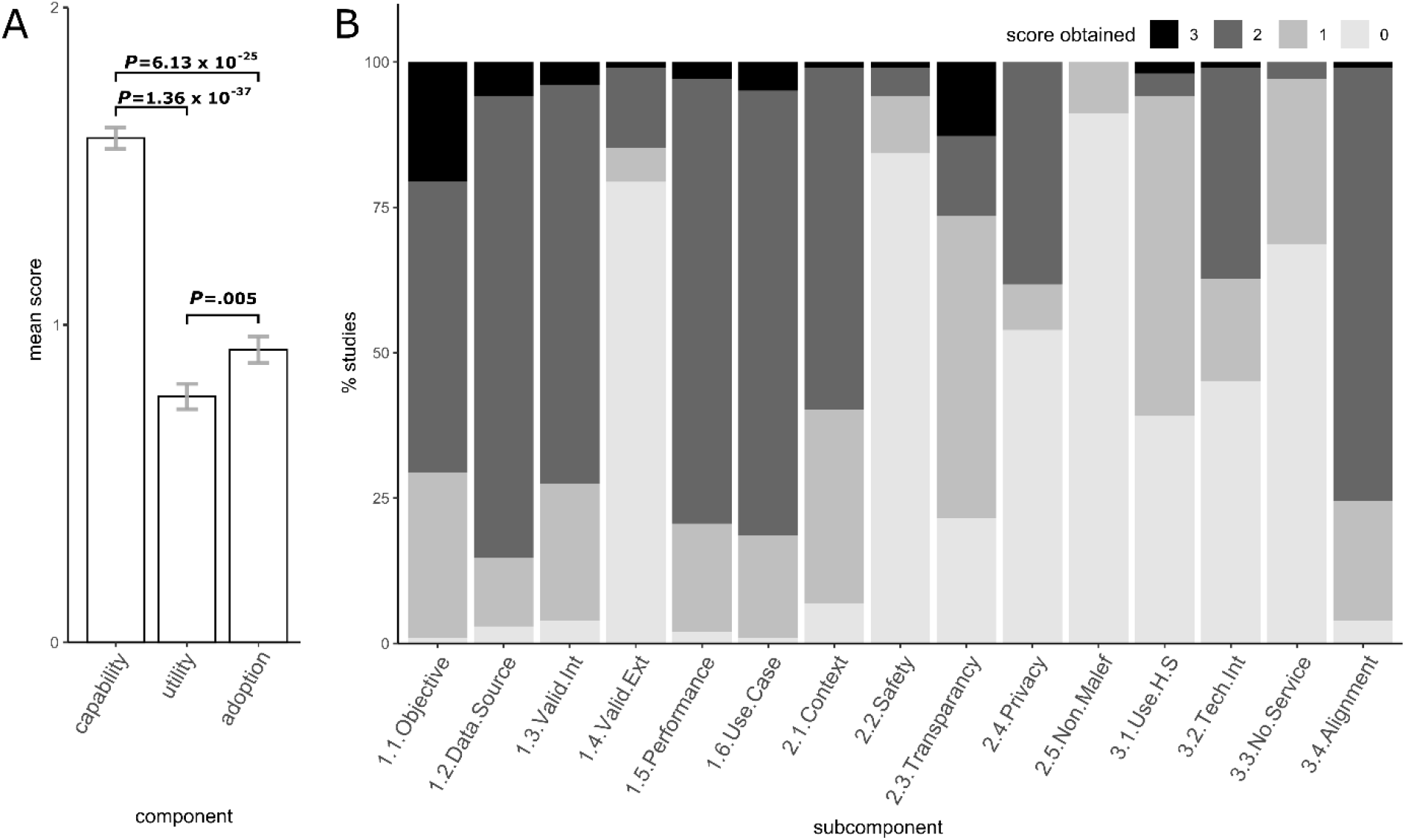
Overall consensus scores obtained by all studies reviewed. A) Average consensus scores for all studies reviewed (error bars = standard error). B) Stacked bar graph showing the distribution of scores for each subcomponent question

We observed moderate positive correlation (R = 0.19-0.43) between most capability component questions (data source vs: internal validation, R = 0.43; external validation, R = 0.20; performance, R = 0.33, and; Use case, R = 0.37. Internal validation vs: performance R = 0.40; use case, R = 0.31. Performance vs use case, R = 0.32), with the exception of the objective of study subcomponent (objective of study vs: data source, R = 0.13; internal validation, R = 0.09; external validation, R = 0.08; performance) (Figure 2). This indicates that if a study scored well in one subcomponent of the capability component, then it was also likely to score well in the other capability subcomponents, with the exception of the objective of study subcomponent. Furthermore, there was also correlation between the subcomponents belonging to the capability component and that of the generalisability and contextualisation (R = 0.19 – 0.31), transparency (R = 0.11 – 0.27) and alignment with domain (R = 0.13 – 0.40) subcomponents, as well as our data extraction question 9 (method of machine learning used) (R = 0.11 – 0.24) (Figure 2). There was also significant, moderate correlation between most adoption component questions (R = 0.18 – 0.42), with the exception of the alignment with domain subcomponent (R = 0.04 – 0.26) (Figure 2). A significant negative correlation is observed between a countries GDP and Imaging studies (R = -0.30), indicating that high GDP countries were less likely to do imaging studies than those countries with middle GDP. The negative correlation between the audience (clinical or methodological) and number of services (R = -0.36) is indicative that methodological studies were less likely to be associated with numerous services than clinical studies. Code availability was inversely correlated with Transparency (R = -0.36), as expected (open source was one of the assessment conditions).

**Figure 2.**
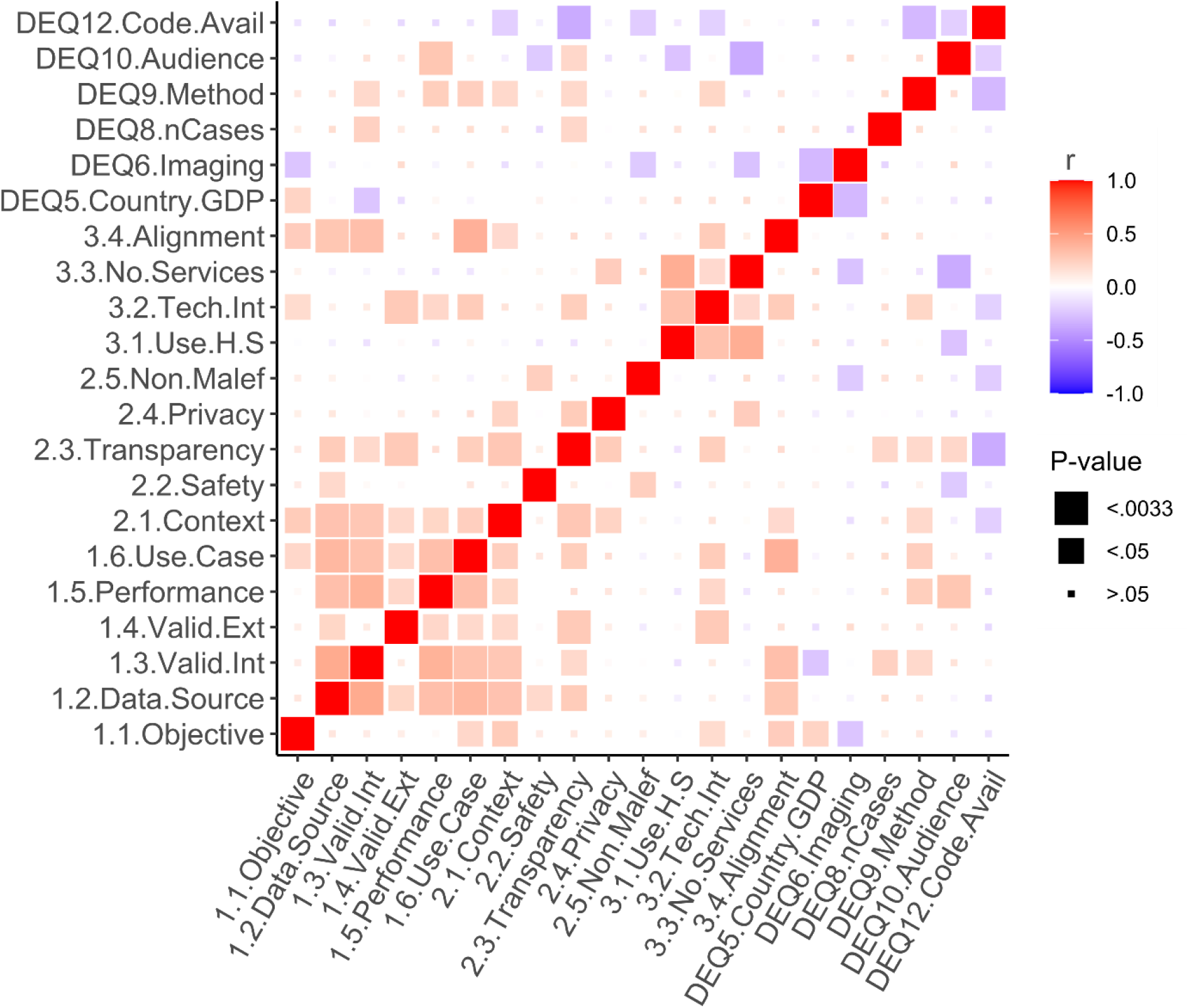
Correlation heatmap showing the strength of correlation between all subcomponents and select data extraction questions. Strength of correlation, as determined by Fisher’s test, is shown by colour with size of squares representing level of significance.

### AI study characteristics

The associations between the AI algorithms used in the studies and TEHAI scores are shown in Figure 3. Deep Learning (including Convolutional Neural Network or CNN for short) was the most frequent machine learning model (54/102 studies) followed by Classic methods (14/102 studies, comprising primarily Linear and Logistic Regression models) and standard Machine Learning (9/102 studies, comprising primarily Random Forest (RF) and Support Vector Machine (SVM) algorithms) (Figure 3A). In 20% of studies, multiple types of algorithms were used. At the component level, Deep learning and Machine learning scored better in capability (mean scores = 1.69 ± 0.04 and 1.54 ± 0.12 respectively) and Deep learning was also superior in adoption (mean score = 0.95 ± 0.06) (Figure 3B). This pattern is also evident at the subcomponent level, where classic methods scored the poorest for most questions (mean scores = 0.07 – 1.78), with deep learning scoring significantly higher in numerous subcomponents (mean scores = 0.05 – 1.96) (Figure 3C). These finding reveal that those using deep learning are more likely to include facets into their design that is more likely to ensure their work will be integrated into practise.

**Figure 3.**
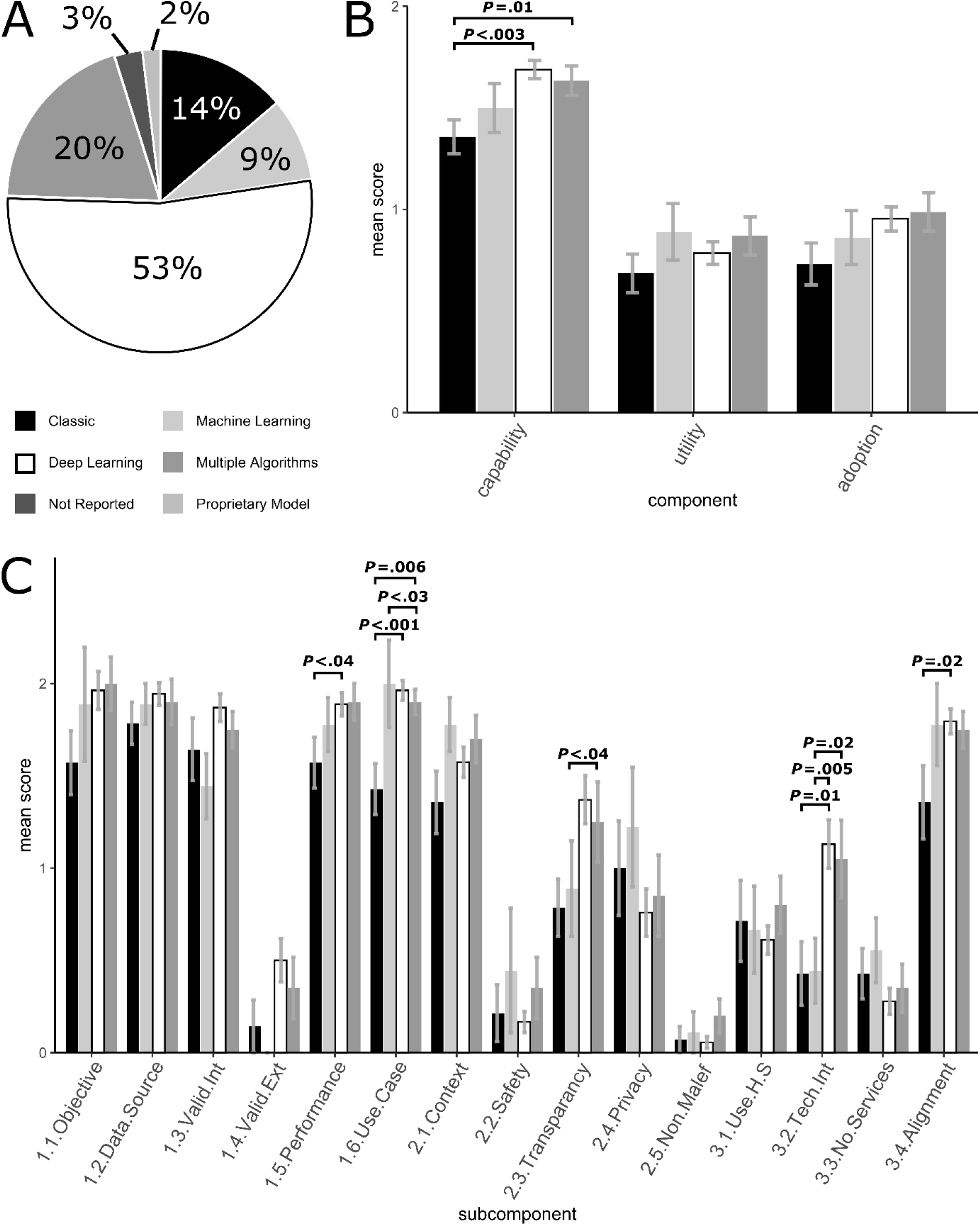
Methods used by the various studies to achieve end-points. A) percentage of studies using specific methods. As the field of potential algorithms is diverse, we created broad categories to make the pie chart readable and to provide an overview of the most prevalent types of algorithms. Classic methods included linear and logistic regression models and the machine learning category comprised a heterogeneous mix of established non-linear algorithms such as random forest and support vector machine. The deep learning category included mostly convoluted neural networks and represented more recent neural network techniques developed for big data. B) “component” scores for the four main methods utilised in the studies. C) “subcomponent” scores for the four main methods utilised in the studies. Bars show average scores, with error bars equal to standard error. Bold P-values indicate P < .05. Bonferroni corrected significance P=.0033.

Figure 4 contains the results from comparisons between clinical and methodologically focused papers. Methodological studies tended to score higher in the capability component (methodological mean score = 1.63 ± 0.04, clinical mean score = 1.52 ± 0.06), and clinically focused studies tended to score higher in utility (clinical mean score = 0.81 ± 0.07, methodological mean score = 0.75 ± 0.05) and adoption (clinical mean score = 1.03 ± 0.07, methodological mean score = 0.87 ± 0.05) (Figure 4A), particularly in the use in a healthcare setting (clinically focused mean score = 0.90 ± 0.11, methodologically focused mean score = 0.58 ± 0.08, *P* = .037) and number of services (clinically focused mean score = 0.58 ± 0.09, methodologically focused mean score = 0.23 ± 0.06, *P* = 2.39 × 10^−05^) subcomponents. It is important to note that all papers scored poorly in safety (clinically focused mean score = 0.13 ± 0.14, methodologically focused mean score = 0.58 ± 0.05) and non-maleficence (clinically focused mean score = 0.12 ± 0.06, methodologically focused mean score = 0.07 ± 0.03) subcomponents and despite being more integrated into the health system, clinical papers did not score significantly higher scores in these subcomponents (Figure 4A and 4B).

**Figure 4.**
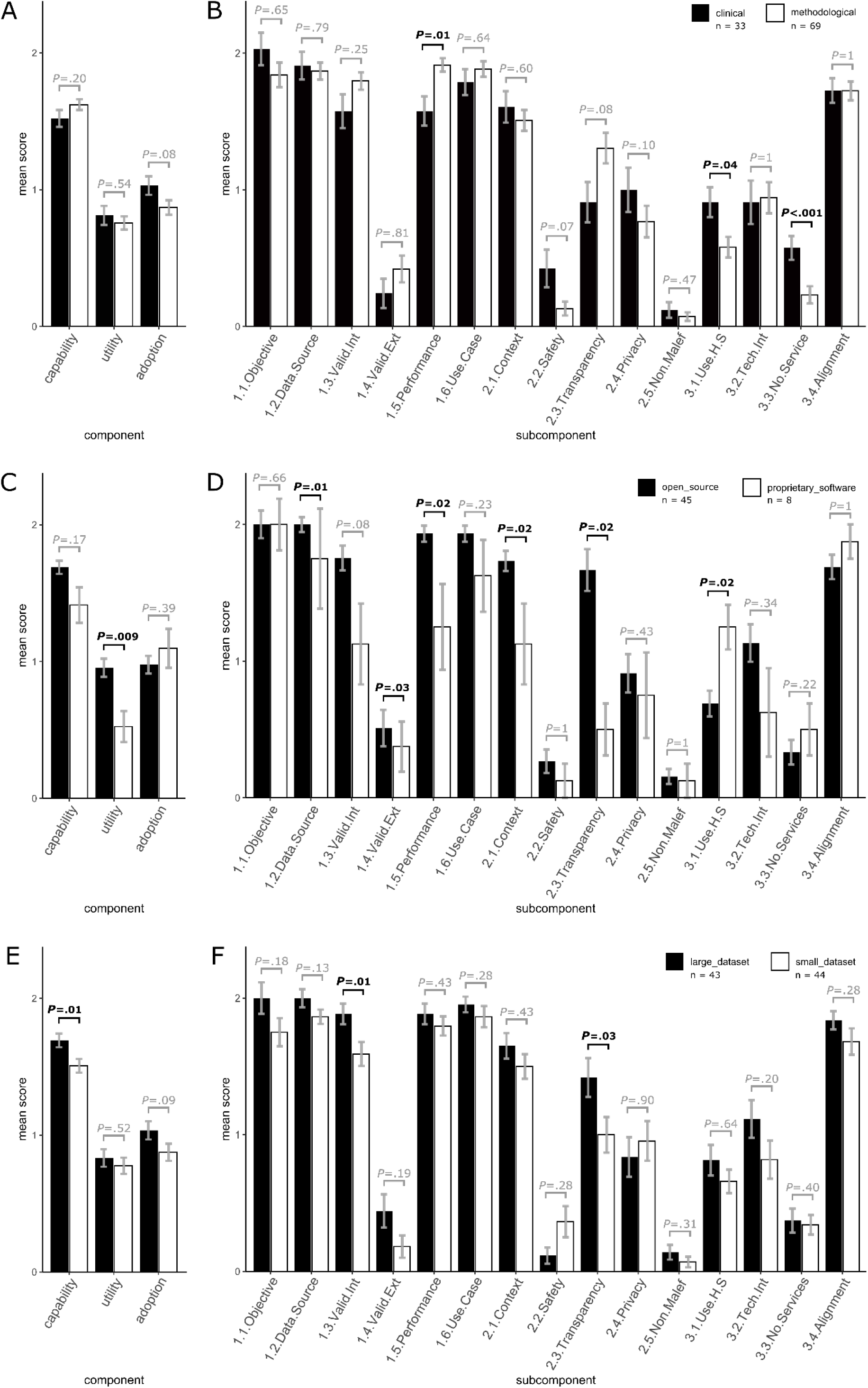
Component and subcomponent scores split into subcategories based on data extraction questions, including: A and B) “intended audience”, C and D) “type of software”, and E and F) “size of dataset”. Bars show average scores, with error bars equal to standard error. Bold P-values indicate P < .05. Bonferroni corrected significance P=.0033.

Close to half of the studies used open source software (n=45), with a small portion (n=8) using proprietary software (with the remaining studies being unclear as to the software availability). There was a tendency for proprietary software to perform better at adoption, particularly in the use in a healthcare setting subcomponent (open source software studies mean score = 0.69 ± 0.09, proprietary software studies mean score = 1.25 ± 0.16, *P* = .02), while papers with open source software tended to score better in utility, including the safety (open source software studies mean score = 0.27 ± 0.09, proprietary software studies mean score = 0.13 ± 0.13, *P* = 1), privacy (open source software studies mean score = 0.91 ± 0.14, proprietary software studies mean score = 0.75 ± 0.31, *P* = .43) and non-maleficence (open source software studies mean score = 0.15 ± 0.05, proprietary software studies mean score = 0.13 ± 0.16, *P* = 1) (Figure 4C and 4D). We also observed a tendency for open source software to score better at transparency (open source software studies mean score = 1.67 ± 0.15, proprietary software studies mean score = 0.5 ± 0.19, *P* = 0.02), which is compatible with the findings from correlation analysis (Figure 2).

Across the studies, the median number of cases was 225 subjects, therefore we allotted studies with a number of cases greater than 225 cases to the large dataset category and those with less than or equal to 225 cases to the small dataset category (Figure 4E and 4F). There was an overall suggestive pattern for the large dataset to score higher than the small dataset, again with the exception of safety, privacy and both scored poorly for non-maleficence.

Countries may have differing capacities to integrate new technologies into their health system and we hypothesized that it would be detectable via GDP. We split the studies into low-, middle- and high-income countries based on classifications as defined by the world bank[23]. There were no countries that published studies that were in the low-income category, with half of the studies originating in middle-income countries and the other half in high-income countries. Interestingly there was no significant difference between components at the multiple testing threshold, however there was a trend suggesting a difference in the adoption component (high income study mean score = 1.0 ± 0.06, medium income study mean score = 0.83 ± 0.06, *P* = 0.04) (Multimedia appendix 4A and Multimedia appendix 4B) and a slight tendency toward middle-income countries to score better at the capability subcomponent questions, particularly objective (high income study mean score = 2.1 ± 0.09, medium income study mean score = 1.76 ± 0.1, *P* = 0.03) and internal validation (high income study mean score = 1.58 ± 0.08, medium income study mean score = 1.88 ± 0.08, *P* = 0.04) (Multimedia appendix 4B).

We found that there were many studies where the authors used AI to analyse images of lungs (e.g. x-rays) of COVID-19 patients and controls to classify them into categories, ultimately producing algorithms that could accurately identify COVID-19 patients from images of their lungs. Thus we classified studies as being imaging (direct image analysis of X-rays or CT scans) or non-imaging (e.g. studies that analysed blood metabolites) and there was a strong trend for non-imaging studies to score higher than imaging studies, this includes the subcomponents of objective (imaging study mean score = 1.79 ± 0.08, non-imaging study mean score = 2.18 ± 0.13, P = 0.02), safety (imaging study mean score = 0.16 ± 0.05, non-imaging study mean score = 0.36 ± 0.14, *P* = 0.015), non-maleficence (imaging study mean score = 0.04 ± 0.02, non-imaging study mean score = 0.18 ± 0.07, *P* = 0.05) and number of services (imaging study mean score = 0.25 ± 0.06, non-imaging study mean score = 0.55 ± 0.11, *P* = 0.02) (Multimedia appendix 4C and D).

## Discussion

Considering the emergence of the COVID-19 pandemic and the flurry of AI models that were developed to address various aspects of the pandemic, we conducted a systematic review of these AI models regarding their likely success at translation. We observed a significant trend for studies to attain high scores for technical capability but low scores for the areas essential for clinical translatability. Specific questions regarding external model validation, safety, non-maleficence and service adoption received failed scores in most studies. Therefore, we identified notable quality gaps in most AI studies of COVID-19 that are likely to have a negative impact on clinical translation.

There have been many claims made of such AI models, including similar or higher accuracy, sensitivity and/or specificity compared with human experts [24–26] and real-time results that have been suggested to lead to improved referral adherence [27] but very few independent studies have tested these claims. In fact, it is suggested that while the AI models have potential, they are generally unsuitable for clinical use and, if deployed prematurely, could lead to undesirable outcomes including stress on both patients, unnecessary intrusive procedures, and the health system and even death due to misdiagnosis [5,7]. Of those studies that examined the utility of COVID-19 AI applications there has not been a comprehensive evaluation of AI in healthcare models encompassing assessment of their intrinsic capabilities, external performance, and adoption in healthcare delivery thus far. It is important for the scientific community and relevant stakeholders to understand how many of these AI models are translational in their value and to what degree. To address this gap, we undertook a comprehensive evaluation of COVID-19 AI models that were developed between December 2019-December 2020. The framework we chose, TEHAI, is a comprehensive evaluation framework developed by a multi-disciplinary international team through a vigorous process of review and consultation, and systematically assesses AI models for their translational value [15]. To select the COVID-19 studies, we conducted a systematic search and after screening 3830 studies, we selected 102 studies for the evaluation. As per TEHAI, the studies were assessed for their capability, utility and adoption aspects and scored using a weighted process.

The scale of the studies we screened (over 3000) and the studies eligible for evaluation (over 900) indicate the level of activity in this area despite the limited time frame selected for the evaluation (2019-2020). The evaluation of the 102 studies while yielding some interesting findings also had a few expected results. Notable was most studies while doing well in the capability component, did not evaluate highly in the utility and adoption components. The latter components assess the ethical, safety and quality and integration with healthcare service aspects of the AI model. However, it is not surprising the AI models scored low in these components, given the expediency required to develop and release these models in a pandemic context. This meant the ethical components were not a priority as you would expect in normal times. It was also not surprising to find that convolutional neural network was the most popular machine learning model as most of the selected studies related to medical imaging analysis (69/102 studies were imaging compared with 33 that were not), where the technique is widely understood and beginning to be applied in some clinical settings [6,28].

While there was a consistent trend for studies with a large dataset to score higher than those with small datasets, there was no significant difference for any sub-component between studies with small versus large datasets was a surprising finding, this indicates that even when studies have collected more data, they advance no further in the utility or adoption fields, should the total number of studies analysed be increased, we would expect the difference between the two datasets to become significant. Regarding imaging vs. non-imaging, we observed that non-imaging studies scored higher in some adoption and utility subcomponents; we suspect this was due to the more clinical nature of the non-imaging research teams, thus the papers focused more on issues important to clinical practice. While there was a tendency for those studies using proprietary software that we expected to be more mature, the authors had not advanced the findings into practice any more than that of open-source algorithm-based studies, again we would expect this difference to become significant if the number of studies scored were to be increased. We also assessed the interpretability of the models as part of the Transparency subcomponent and found that image studies, in particular, included additional visualization to pinpoint the regions that were driving the classification. Further, the scoring studies in each of the TEHAI components evidenced the need for planning in advance for external validation, safety, and integration in health services to ensure the full translatability of AI models in healthcare.

Most of the reviewed studies lacked sufficient considerations for adoption into health care practices (the third TEHAI component), which has implications for the business case for AI applications in health care. Cost of deployment and costs from misclassification from both monetary and patient safety/discomfort perspective can only be assessed if there is pilot data available from actual tests that put new tools into service. Furthermore, critical administrative outcomes such as workload requirements should be considered as early as possible. While we understand that such tests are hard to organize from academic basis, the TEHAI framework can be used as a incentive to move into this direction.

“We note that availability of dedicated datasets and computing resources for training could be a bottleneck for some applications. In this study, we observed multiple instances of transfer learning, which is one solution, however, we will revise the capability section of the TEHAI to make more specific consideration for these issues.” Fair access to AI technology should also be part of a good design. The TEHAI frameworks includes this in the internal validity subcomponent, where we small studies, in particular, struggled with representing a sufficient diversity of individuals. From a translational point of view, we also observed short comings in the contextualization of AI models. Again, since there was limited evidence on service deployment, most studies scored low on fairness simply due to lack of data. We also note that deployment in this case may be hindered by clinical acceptance of the models [11], and we will include this topic in future amendments to the TEHAI framework.

### Limitations

While we undertook a comprehensive evaluation of AI studies unlike previous assessments, our study still has some limitations. Firstly, the period we used to review and select studies was narrow, being just a year. Another limitation is that for practical reasons we randomly chose a subset of 102 studies for evaluation out of the 968 eligible studies. Despite these limitations, we are confident the evaluation process we undertook was rigorous as evidenced by the systematic review of literature, detailed assessment of each of the selected studies and the parallel review and consensus steps.

We recommend caution when generalizing the results from this COVID-19 study to other areas of AI in health care. Firstly, evaluation frameworks that rely on human experts can be sensitive to the selection of the experts (subjectivity). Secondly, scoring variation may arise from the nature of the clinical problem rather than the AI solution per se, thus TEHAI results from different fields may not be directly comparable. Thirdly, we intentionally excluded discovery studies aimed at new biology or novel treatments as those would have been too early in the translation pipeline to have a meaningful evaluation. Fourthly, there is also capacity for significant heterogeneity of clinical domains may confound the evaluation results and may prevent comparisons of studies (here we made effort to pre-select studies that were comparable). Lastly, the TEHAI framework was designed to be widely applicable, which means that stakeholders with specific subjective requirements may need to adapt their interpretations accordingly.

We acknowledge the rapid progress in AI algorithms that may make some of the evaluation aspects obsolete over time, however, we also emphasize that two out of the three TEHAI components are not related to AI itself, but to the ways AI interacts with the requirements of clinical practice and health care processes. Therefore, we expect that the translatability observations from this study will have longevity.

## Conclusions

AI in healthcare has a translatability challenge as evidenced by our evaluation study. By assessing 102 AI studies for their capability, utility, and adoption aspects we uncovered translational gaps in many of these studies. Our study highlights the need to plan for translational aspects very early in the AI development cycle. The evaluation framework we used and the findings from its application will inform developers, researchers, clinicians, authorities, and other stakeholders to develop and deploy more translatable AI models in healthcare.

## Supporting information

Multimedia Appendix 1

Multimedia Appendix 2

Multimedia Appendix 3

Multimedia Appendix 4

## Data Availability

All data produced in the present study are available upon reasonable request to the authors

## Acknowledgments

B.K extracted appropriate studies from databases. A.C. assigned studies to reviewers. All authors were involved in the scoring process. A.C. carried out all analysis and generated figures. A.C., S.R., S.A. and V-P.M. drafted the manuscript. All authors provided feedback and edits for the final manuscript.

## Multimedia Appendix

Multimedia Appendix 1. PRISMA Flow Diagram.

Multimedia Appendix 2. Search Strategies.

Multimedia Appendix 3. Evaluation and Scoring Questions.

Multimedia Appendix 4. Component and subcomponent scores split into subcategories based on data extraction questions, including: A and B) “country GDP”, C and D) “imaging/non-imaging” based study. Bars show average scores, with error bars equal to standard error. Bold P-values indicate P < .05. Bonferroni corrected significance P=.0033.

## Abbreviations

AI: Artificial Intelligence
COVID-19: Corona Virus Disease 2019
SE: Standard Error
TEHAI: ‘Translational Evaluation of Healthcare Artificial Intelligence

