## Supplementary material for "Application of comprehensive evaluation framework to Coronavirus Disease 19 studies: A systematic review of translational aspects of artificial intelligence in health care": Multimedia Appendix 1

**Identification of studies via databases and registers**

Records removed before screening:

Duplicate records removed (n = 1446)

Records identified from databases:

MEDLINE (n = 2389)

Embase (n = 1897)

iSearch COVID-19 portfolio (n = 990)

Total (n = 5276)

**Identification**

Records screened

(n = 3830)

Records excluded

(n = 2862)

Reports relevant and eligible for evaluation

(n = 968)

Reports not randomly selected for evaluation (n = 845)

**Screening**

Reports excluded from evaluation:

Did not address a healthcare problem (n = 11 )

Did not use artificial intelligence (n = 7)

Wrong study type (n = 2)

Reports randomly selected for evaluation (n = 123)

Studies included in evaluation

(n = 102)

**Included**
