## Supplementary material for "Application of comprehensive evaluation framework to Coronavirus Disease 19 studies: A systematic review of translational aspects of artificial intelligence in health care": Multimedia Appendix 2

### **Search Strategies for MEDLINE, Embase and NIH iSearch databases**

**MEDLINE(R) and In-Process, In-Data-Review & Other Non-Indexed Citations searched via OvidSP**

1. (coronavirus* or corona virus* or ncov* or sars-cov* or sarscov* or sars-coronavirus* or severe acute respiratory syndrome coronavirus* or 2019-ncov or ncov19 or ncov-19 or 2019-novel or sars-cov2 or sars-cov-2 or sarscov2 or sarscov-2 or sars-coronavirus2 or sars-coronavirus-2 or sars-like coronavirus* or coronavirus-19 or covid19 or covid-19 or covid 19 or covid 2019).ab,ti.
2. (exp Coronavirus/ or Coronavirus Infections/ or Severe Acute Respiratory Syndrome/)
3. (1 or 2)
4. (AI or artificial intelligence* or deep learning or machine learning or neural network* or risk model*).ab,ti.
5. (Algorithms/ or exp Artificial Intelligence/)
6. (4 or 5)
7. (3 and 6)
8. 20191201:20301231.(dt).
9. (7 and 8)

**Embase searched via Embase.com**

#1. (coronavirus* OR “corona virus*” OR ncov* OR sars-cov* OR sarscov* OR sars-coronavirus* OR “severe acute respiratory syndrome coronavirus*” OR 2019-ncov OR ncov19 OR ncov-19 OR 2019-novel OR sars-cov2 OR sars-cov-2 OR sarscov2 OR sarscov-2 OR sars-coronavirus2 OR sars-coronavirus-2 OR “sars-like coronavirus*” OR coronavirus-19 OR covid19 OR covid-19 OR “covid 2019”):ab,ti
#2. ('coronavirinae'/exp OR 'coronavirus infection'/exp)
#3. (#1 OR #2)
#4. (AI OR “artificial intelligence*” OR “deep learning” OR “machine learning” OR “neural network*” OR “risk model*”):ab,ti
#5. ('artificial intelligence'/exp or 'machine learning'/exp)
#6. (#4 OR #5)
#7. (#3 AND #6)
#8. [1-12-2019]/sd NOT [1-1-2031]/sd
#9. (#7 AND #8)

**NIH iSearch COVID-19 Portfolio searched via https://icite.od.nih.gov/covid19 with searchable fields set to title and abstract only**

(AI OR “artificial intelligence” OR “deep learning” OR “machine learning” OR “neural network” OR “neural networks” OR “neural networking” OR “risk model” OR “risk models” OR “risk modelling”)
