## Supplementary material for "Application of comprehensive evaluation framework to Coronavirus Disease 19 studies: A systematic review of translational aspects of artificial intelligence in health care": Multimedia Appendix 3

### **Evaluation Framework Checklist and Scoring Sheet**

**1. Capability**

**1.1 Objective of study**

This subcomponent assesses whether the study has a clear objective i.e., stated contribution to a specific healthcare field.

This subcomponent is scored on a scale of how clearly the objective is articulated.

1.
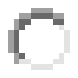
Not applicable to type of study (NA)
2.
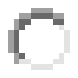
Not reported, considered or acknowledged (0)
3.
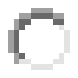
The objective of the study is articulated (1)
4.
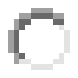
The objective of the study and why the study is being conducted are clearly articulated (2)
5.
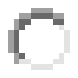
The objective of the study and why the study is being conducted and how it adds to the body of knowledge in the domain are clearly articulated (3)

Clear above selection

**1.2 Dataset Source and Integrity**

An AI model is only as good as the data it was derived from. If the training data does not reflect the intended purpose, the model predictions are likely to be useless or even harmful.

This subcomponent evaluates the source of the data and the integrity of datasets used for training and testing the AI model including an appraisal of the representation of the target population in the data, coverage, accuracy and consistency of data collection processes and transparency of datasets.

This subcomponent is scored on a scale of how well the dataset is described, how well the datasets fits with the ultimate objective and use case and how credible/reliable the data source is.

1.
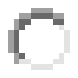
Not applicable to type of study (NA)
2.
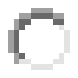
Not reported, considered or acknowledged (0)
3.
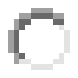
Non-representative training set, low quality or poorly defined collection protocol, dataset biased due to conflict of interests (1)
4.
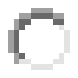
Training set is a sample from intended target population, appropriate data collection protocol, conflicts of interests not likely to drive conclusions (2)
5.
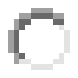
Diverse training set of excellent coverage of all affected population segments, high quality and comprehensive data collection protocol, highly reputable and diverse data creators with no hidden agenda (3)

Clear above selection

**1.3 Internal Validity**

An internally valid model will be able to predict health outcomes reliably and accurately within a pre-defined set of data resources that were used wholly or partially when training the model. This includes the classical concept of goodness-of-fit, but also cross-validation schemes that derive training and tests sets from the same sources of data.

Scoring is based on the size of the training data set with respect to the health care challenge, the diversity of the data to ensure good modelling coverage, and whether the statistical performance of the model (e.g. classification) is high enough to satisfy the requirements of clinical usefulness.

1.
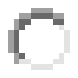
Not applicable to type of study (NA)
2.
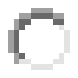
Not reported, considered or acknowledged (0)
3.
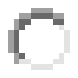
Small internal datasets, low statistical power, poorly defined or low-quality prediction target, inappropriate study design (1)
4.
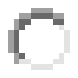
Adequate statistical power, sufficient data quality and accuracy, appropriate study design (2)
5.
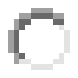
Extensive and representative internal datasets, high quality of measurements, careful consideration for confounders, gold standard prediction target (3)

Clear above selection

**1.4 External Validity**

To qualify as external validation, we require that the external data used to assess model performance must come from substantially distinct external source that did NOT contribute any data towards model training.

Examples of external data sources include independent hospitals, institutions or research groups that were not part of the model construction team or a substantial temporal difference between the training and validation data collections.

The scoring is based on the size and diversity of the external data (if any) and how well the external data characteristics fit with the intended care recipients under the study objective.

1.
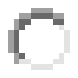
Not applicable to type of study (NA)
2.
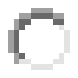
Not reported, considered or acknowledged (0)
3.
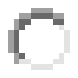
Mismatched internal and external datasets (samples of different populations, data collected differently, outcomes defined differently), small or low-quality validation set (1)
4.
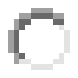
Compatible internal and external datasets, sufficient statistical power, validation data from a different independent source from any training samples (2)
5.
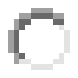
Extensive and multiple external validation datasets from diverse sources, excellent coverage of intended target population and real-world practice (3)

Clear above selection

**1.5 Performance Metrics**

Performance metrics refers to mathematical formulas that are used for assessing how well an AI model predicts clinical or other health outcomes from the data. If the metrics are chosen poorly, it is not possible to assess the accuracy of the models reliably. Furthermore, specific metrics have biases, which means the use of multiple metrics is recommended for robust conclusions.

This subcomponent examines whether performance measures relevant to the model and the results stated in the study are presented. These performance metrics can be classification or regression or qualitative metrics.

This subcomponent is scored on a scale of how well the performance metrics fit the study and how reliable they are likely to be considering the nature of the health care challenge.

1.
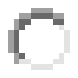
Not applicable to type of study (NA)
2.
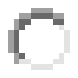
Not reported, considered or acknowledged (0)
3.
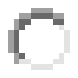
Only one formula used, uncertain accuracy of the performance measures, no replicates for cross-validation or similar framework (1)
4.
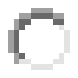
Multiple metrics applied including those relevant for clinical practice, replicates or other techniques applied to consider reliability of performance measures (2)
5.
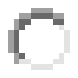
Extensive analyses and benchmarking of relevant performance metrics across multiple datasets, careful experiments to verify the accuracy of performance values, mitigation of potential confounding factors (3)

Clear above selection

**1.6 Use Case**

This subcomponent is seeking justification for the use of AI for the study as opposed to other statistical or analytical methods. This tests if the study has considered the relevance and fit of the AI to the particular healthcare domain it is being applied to.

This subcomponent is scored on a scale of how well the use case is stated.

1.
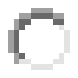
Not applicable to type of study (NA)
2.
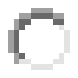
Not reported, considered or acknowledged (0)
3.
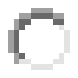
The case for AI use is vaguely formulated or not justified or inappropriate for the dataset, study design or objective (1)
4.
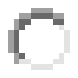
The case for AI use is reasonable (against other alternatives) given the study design and results, and relevant for practice (2)
5.
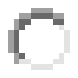
The case for AI use is clearly formulated or justified and appropriate for the dataset, study design or objective; AI is the best foreseeable solution to the health care challenge (3)

Clear above selection

**2. Utility**

**2.1 Generalisability and Contextualisation**

The context of an AI application is defined here as the match between the model performance, expected features, characteristics of the training data and the overall objective.

In particular, biases or exacerbation of disparities due to underrepresentation or inappropriate representation due to the availability of datasets used both in training and validation can have an adverse effect on the real-world utility of an AI model.

This subcomponent is scored based on how well it is expected to perform on the specific groups of people it is most intended for.

1.
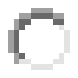
Not applicable to type of study (NA)
2.
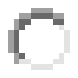
Not reported, considered or acknowledged (0)
3.
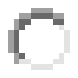
Limited diversity of people or insufficient clinical description to confirm if representative of intended care recipients (1)
4.
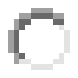
Representative of intended care recipients, appropriate eligibility criteria and baseline characteristics for practical applications (2)
5.
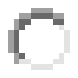
Real-world setting of diverse care patients, excellent coverage of multiple ethnic, socioeconomic and identity groups (3)

Clear above selection

**2.2 Safety and Quality Issues**

It is critical that AI models being deployed in healthcare, especially in clinical environments, are assessed for their safety and quality. Appropriate consideration should be paid to the presence of ongoing monitoring mechanisms in the study, such as adequate clinical governance that will provide a systematic approach to maintaining and improving the safety and quality of care within a healthcare setting.

This subcomponent is scored based on the strength of the S&Q process and how likely it is to ensure S&Q when AI is applied in the real-world.

1.
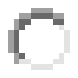
Not applicable to type of study (NA)
2.
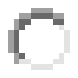
Not reported, considered or acknowledged (0)
3.
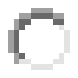
Safety measures and quality controls presented are likely to be inadequate (1)
4.
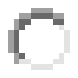
Reasonable steps taken to adopt safety and quality processes but an ongoing sustainable governance framework is not in place (2)
5.
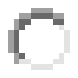
Careful consideration and testing of possible adverse impacts, continuous safety and quality monitoring mechanisms are in place and active (3)

Clear above selection

**2.3 Transparency**

This subcomponent assesses the extent to which model functionality and architecture is described in the study and the extent to which decisions reached by the algorithm are understandable (i.e., black box or interpretable).

Important elements are the overall model structure, the individual model components, the learning algorithm, and how the specific solution is reached by the algorithm. Open source? Public datasets?

This subcomponent is scored on a scale of how transparent, interpretable and reproducible the AI models are, given the information available.

1.
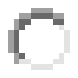
Not applicable to type of study (NA)
2.
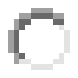
Not reported, considered or acknowledged (0)
3.
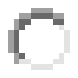
Basic algorithm concept published in a peer-reviewed journal or a technical report, sufficient technical description to reproduce essential components of the pipeline (1)
4.
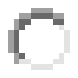
Algorithm is published and source code or software tools are available for inspection, sufficient statistical analyses to describe the patterns in the data that are likely driving the prediction outcomes (2)
5.
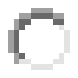
Fully open-source project with public access to training data; the process by which the algorithm derives predictions from data is analysed statistically, carefully documented and visualised/presented in ways that are easy to interpret by humans (3)

Clear above selection

**2.4 Privacy**

This sub-component covers personal privacy, data protection and security.

This subcomponent is ethically relevant to the concept of autonomy/self-determination, the right to control access to and use of personal information, and the consent processes used to authorize data uses.

This subcomponent is scored on the extent of consideration of privacy aspects including consent by study subjects, the strength of data security and data life cycle throughout the study itself and consideration for future protection if deployed in the real-world.

1.
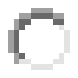
Not applicable to type of study (NA)
2.
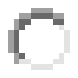
Not reported, considered or acknowledged (0)
3.
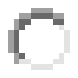
Informed consent when applicable, personal and sensitive data protected from unauthorized access (1)
4.
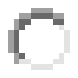
Anonymised research data, study protocol conforms to international standard on privacy/ethics, approved by local ethics committee, informed consent when applicable (2)
5.
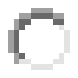
Privacy by design, peer-reviewed study protocol approved by ethics committee and satisfies the highest international standards, facility to opt-out at any time, past and future data securely stored with database usage, disposal plan in place (3)

Clear above selection

**2.5. Non-Maleficence**

This sub-component refers to the identification of actual and potential harms caused by the AI and actions to avoid foreseeable or unintentional harms.

Harms to individuals may be physical, psychological, emotional, economic. Harms may affect systems/organizations, infrastructure and social wellbeing.

This subcomponent is scored on the extent to which potential harms of the AI are identified, quantified and the measures taken to avoid harms and reduce risk.

1.
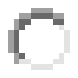
Not applicable to type of study (NA)
2.
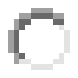
Not reported, considered or acknowledged (0)
3.
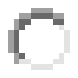
Potential harms are acknowledged (e.g., misdiagnoses, bodily harms, discrimination, loss of trust, loss of skills) but effective mitigation not presented (1)
4.
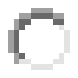
Potential harms are discussed (see above) and specific actions to minimise harms are presented (2)
5.
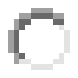
Systematic approach to minimise harms, demonstrated effectiveness of risk management, independent and appropriate audits and governance (3)

Clear above selection

**3. Adoption**

**3.1 Use in a Healthcare Setting**

As discussed earlier, many AI models have been developed in controlled environments or in-silico, but there is a need to assess for evidence of use in real world environments and integration of new AI models with existing information systems. This subcomponent is scored according to the extent to which the model has been adopted by and integrated into ‘real world’ healthcare services i.e., healthcare settings beyond the test site.

1.
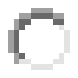
Not applicable to type of study (NA)
2.
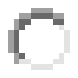
Not reported, considered or acknowledged (0)
3.
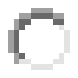
Vague or insufficient description of practical integration into hospital or other care environments beyond the test site (1)
4.
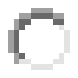
Detailed and convincing plan to integrate methodology into specific clinical/administrative workflows beyond the test site (2)
5.
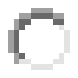
Practical demonstration of how method is integrated into workflows across institutes, governance oversight clearly defined and in place beyond the test site (3)

Clear above selection

**3.2 Technical Integration**

This subcomponent evaluates how well the models integrate with existing clinical/administrative workflows outside of the development setting, and their performance in such situations. In addition, the subcomponent includes reporting of integration even if the model performs poorly.

This subcomponent is scored on a scale of how well the integration aspects of the model are anticipated and if specific steps to facilitate practical integration have been taken.

1.
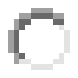
Not applicable to type of study (NA)
2.
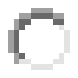
Not reported, considered or acknowledged (0)
3.
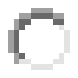
Insufficient model performance for purpose, poor fit, presence of artefacts (1)
4.
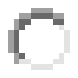
Solid evidence for sufficient model performance and accuracy for purpose across multiple metrics (2)
5.
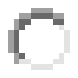
Extensive evidence for sufficient and stable model performance for purpose across multiple settings and datasets, highly likely or demonstrated performance in intended real-world environment (3)

**3.3 Number of Services**

Many AI in healthcare studies are based on single site use without evidence of wider testing or validation. In this subcomponent, we review reporting of wider use.

This subcomponent is scored on a scale of how well the use of the model across multiple healthcare organisations is described.

1.
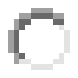
Not applicable to type of study (NA)
2.
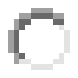
Not reported, considered or acknowledged (0)
3.
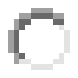
Deployed or piloted in one healthcare service or entity (1)
4.
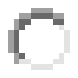
Established long-term deployment in a single large healthcare service or multiple national deployments across different services (2)
5.
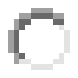
International deployment across multiple healthcare services and entities with a demonstrated record of success (3)

**3.4 Alignment with Domain**

This category considers how well the study described the alignment and relevance of the AI model and its results to the particular healthcare domain and its likely long-term acceptance.

In other words, the model is assessing the benefits of the AI model to the particular medical domain the model is being applied to. This again relates to the translational aspects of the AI model.

This subcomponent is scored on a scale of how well the benefits of the AI model to the medical domain are articulated.

1.
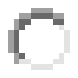
Not applicable to type of study (NA)
2.
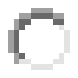
Not reported, considered or acknowledged (0)
3.
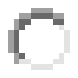
Poor justification for clinical benefit due to lack of societal need, clinical evidence or scientific rationale (1)
4.
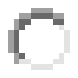
Sound justification for added clinical benefit from AI, good methodological fit between chosen approach and problem to be solved (2)
5.
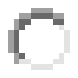
Objective is a direct response to pressing medical need, high likelihood of transformative impact, solution difficult without AI (3)
