## Supplementary figures and images for "Application of comprehensive evaluation framework to Coronavirus Disease 19 studies: A systematic review of translational aspects of artificial intelligence in health care"

### Multimedia Appendix 4

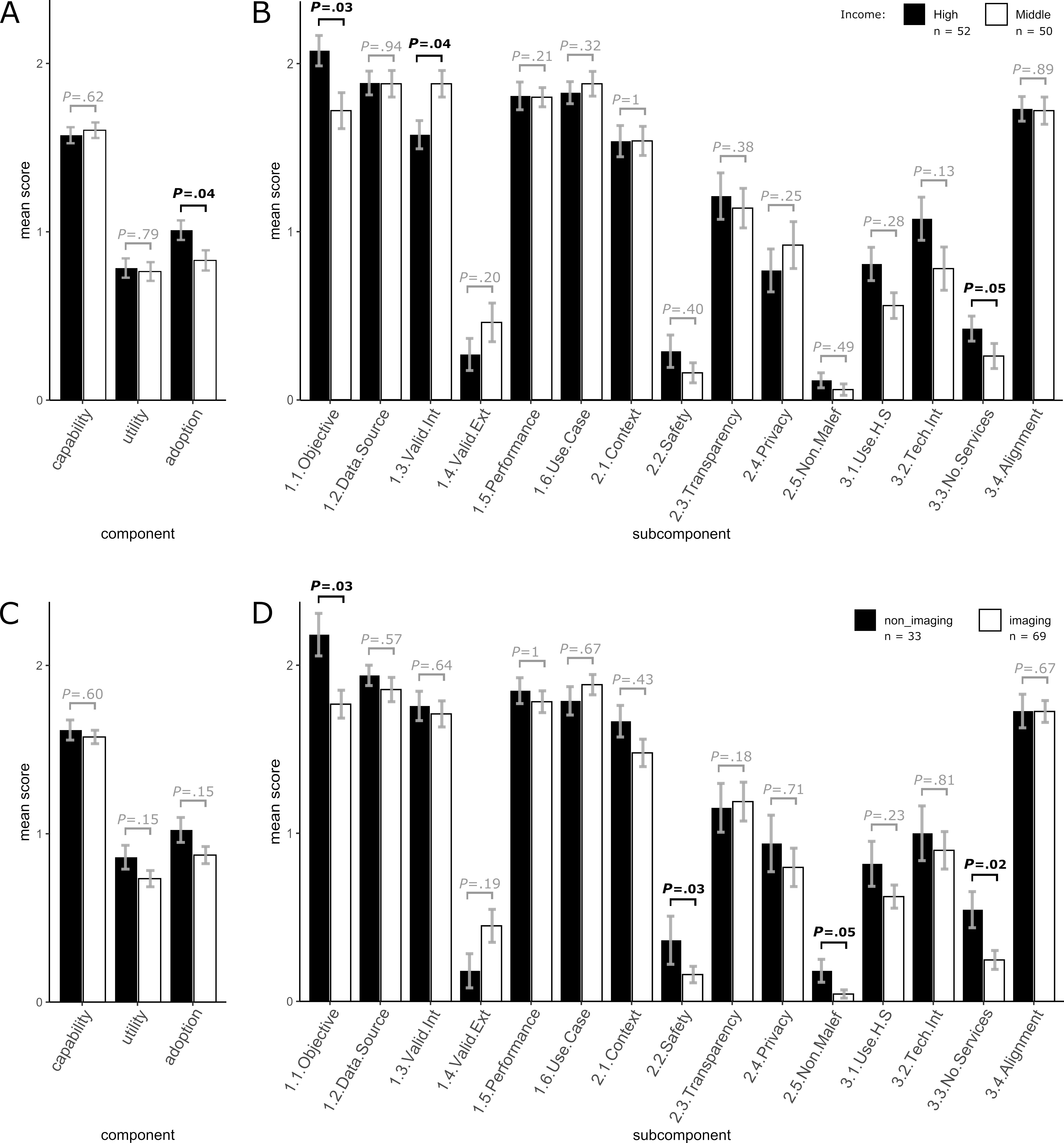
